# Occupational differences in SARS-CoV-2 infection: Analysis of the UK ONS Coronavirus (COVID-19) Infection Survey

**DOI:** 10.1101/2022.04.28.22273177

**Authors:** Sarah Rhodes, Jack Wilkinson, Neil Pearce, Will Mueller, Mark Cherrie, Katie Stocking, Matthew Gittins, Srinivasa Vittal Katikireddi, Martie Van Tongeren

**Affiliations:** Centre for Biostatistics, School of Health Sciences, Faculty of Biology, Medicine and Health, School of Health Sciences, The University of Manchester, Manchester, Manchester, UK; Epidemiology and Population Health, London School of Hygiene & Tropical Medicine, London, UK; Institute of Occupational Medicine, Edinburgh, UK; MRC/CSO Social and Public Health Sciences Unit, University of Glasgow, Glasgow, UK; Centre for Occupational and Environmental Health, School of Health Sciences, Faculty of Biology, Medicine and Health, University of Manchester, Manchester, Greater Manchester, UK

## Abstract

**Background:** Considerable concern remains about how occupational SARS-CoV-2 risk has evolved during the COVID-19 pandemic. We aimed to ascertain which occupations had the greatest risk of SARS-CoV-2 infection and explore how relative differences varied over the pandemic.

**Methods:** Analysis of cohort data from the UK Office of National Statistics Coronavirus (COVID-19) Infection Survey from April 2020 to November 2021. This survey is designed to be representative of the UK population and uses regular PCR testing. Cox and multilevel logistic regression to compare SARS-CoV-2 infection between occupational/sector groups, overall and by four time periods with interactions, adjusted for age, sex, ethnicity, deprivation, region, household size, urban/rural neighbourhood and current health conditions.

**Results:** Based on 3,910,311 observations from 312,304 working age adults, elevated risks of infection can be seen overall for social care (HR 1.14; 95% CI 1.04 to 1.24), education (HR 1.31; 95% CI 1.23 to 1.39), bus and coach drivers (1.43; 95% CI 1.03 to 1.97) and police and protective services (HR 1.45; 95% CI 1.29 to 1.62) when compared to non-essential workers. By time period, relative differences were more pronounced early in the pandemic. For healthcare elevated odds in the early waves switched to a reduction in the later stages. Education saw raises after the initial lockdown and this has persisted. Adjustment for covariates made very little difference to effect estimates.

**Conclusions:** Elevated risks among healthcare workers have diminished over time but education workers have had persistently higher risks. Long-term mitigation measures in certain workplaces may be warranted.

**What is already known on this topic:** Some occupational groups have observed increased rates of disease and mortality relating to COVID-19.

**What this study adds:** Relative differences between occupational groups have varied during different stages of the COVID-19 pandemic with risks for healthcare workers diminishing over time and workers in the education sector seeing persistent elevated risks.

**How this study might affect research, practice or policy:** Increased long term mitigation such as ventilation should be considered in sectors with a persistent elevated risk. It is important for workplace policy to be responsive to evolving pandemic risks.

## Background

The need to protect workers from COVID-19 is an ongoing issue and is likely to be so for some time. There is much debate around the degree to which SARS-CoV-2 transmission occurs in the workplace and which occupations are most affected, with calls for COVID-19and subsequent long term symptoms and disability (long-COVID) to be classified as an occupational disease (1, 2). Hence, it is important that we better understand occupational risks in order to inform policy and practice. Risk of COVID-19 disease in the workplace will be a consequence of exposure to SARS-CoV-2 virus; workplace factors known to be related to exposure include ventilation, ability to social distance and number of daily contacts (3, 4).

Several studies have found increased risks of infection and mortality from COVID-19 amongst healthcare workers (5-8) and those working in public facing transport occupations when compared to non-essential or other workers. Other studies have not found an increased risk (9) or suggest it varies by type of worker and/or stage of the pandemic (10-16).

Other non-healthcare occupations considered to have high exposure to SARS-CoV-2 include police and protective services, education workers, social care workers, office workers and construction workers (17). The evidence for whether this expected exposure translates to increased infection and/or mortality is varied (7, 10, 16, 18-20).

We analysed data from the Office of National Statistics (ONS) Coronavirus (COVID-19) Infection Survey (CIS), with the following objectives:

i. to ascertain whether occupation is associated with SARS-CoV-2 infection;
ii. to ascertain working in which occupations leads to the greatest increased risk of infection from SARS-CoV-2;
iii. to ascertain to what extent increased risks are explained by confounders; and
iv. to explore how relative differences varied in different time periods during the COVID-19 pandemic.

## Methods

### Dataset

The CIS is a randomly sampled repeated cross-sectional household survey with serial sampling designed to be representative of the UK population. The design has been reported in detail elsewhere (21, 22). Each visit included a baseline or follow up survey and a PCR test; participants were followed up regardless of COVID-19 or isolation status. We used data relating to all survey visits from April 2020 to Nov 2021.

### Data analysis

All analyses were restricted to individuals aged 20-64 years at their first ONS visit. Initially we looked at the relationship between occupation and the number of participants with at least one positive PCR test result using basic frequency tables reported as n (%). In all cases the denominator used was the number of individuals aged 20-64 who were active in the CIS during the time period of interest. All infection variables can be considered binary, relating to the number of participants with at least one positive result. Only positive results obtained as part of the ONS survey (and not self-reported results reported between visits) were used.

We analysed the data using time-varying Cox regression, which produces estimates of Hazard Ratios (for a first positive PCR test within the survey) with 95% confidence intervals. Time was measured for each individual from the date of the first survey (April 2020), with individuals censored at their last available follow up.

To explore how relative differences varied over time, the data were divided into four time periods (Table 1), and analysed using a logistic multi-level model with a random intercept for individual. In this analysis, ‘infection within the time period’ was the dependent variable; multiple infections per person were considered (negative test required between subsequent infections). Marginal odds ratios were calculated post-estimation to allow us to report the difference in odds compared to non-essential workers for each time period.

**Table 1.** Details of 4 time periods

| Date | Name | Description |
| --- | --- | --- |
| 1<sup>st</sup> April 2020 to 30<sup>th</sup> Sept 2020 | T1 | End of wave 1. Lifting of national lockdown in June followed by series of local lockdowns. |
| 1<sup>st</sup> Oct 2020 to 28<sup>th</sup> Feb 2021 | T2 | Wave 2. Two periods of national lockdown and the tier system. |
| 1<sup>st</sup> March 2021 to 31<sup>st</sup> May 2021 | T3 | Gradual easing of restrictions. |
| 1<sup>st</sup> June 2021 to 31<sup>st</sup> Oct 2021 | T4 | Wave 3. End of all restrictions. |

We examined other analyses in this area (23) and the Directed-Acyclic Graphs (DAGs) used (24) to derive a plausible DAG on which to base our adjustment set (SX). An interactive version can be accessed at http://dagitty.net/dags.html?id=5J_TeK, which we used to explore implications. We consider our DAG to be suitable to answer questions about the ‘acute’ effect of workplace attendance during the pandemic. Accordingly, we do not represent effects of occupation on health, SES, or living conditions in the DAG, because these are affected by extended tenure in an occupation, rather than by acute exposure. Given our interest in short-term, rather than long-term occupational effects on infection, we treat our SX variables as confounders, rather than mediators. We included ‘non-workplace activities’ in sensitivity analysis only as we had limited data and concerns over measurement error.

All of our analyses followed the sequence in Table 2. Age was included in the model as quintiles, and other variables utilised the categories seen in Table 3. No adjustment was made for multiple participants from the same household (due to model convergence issues) and no weighting was used (due to available weights being cross sectional rather than longitudinal). Coefficient plots were used to compare the occupational effects across the different models. We focus on model 3 in the text which is intended to reflect the direct effect of work-related risks.

**Table 2.** Table of adjustment set by model

| Model number | Covariates |
| --- | --- |
| 1 | Age, sex |
| 2 | Age, sex, ethnic group, IMD, region |
| 3 | Age, sex, ethnic group, IMD, region, household size, urban or rural location, presence of health condition. |

**Table 3.**
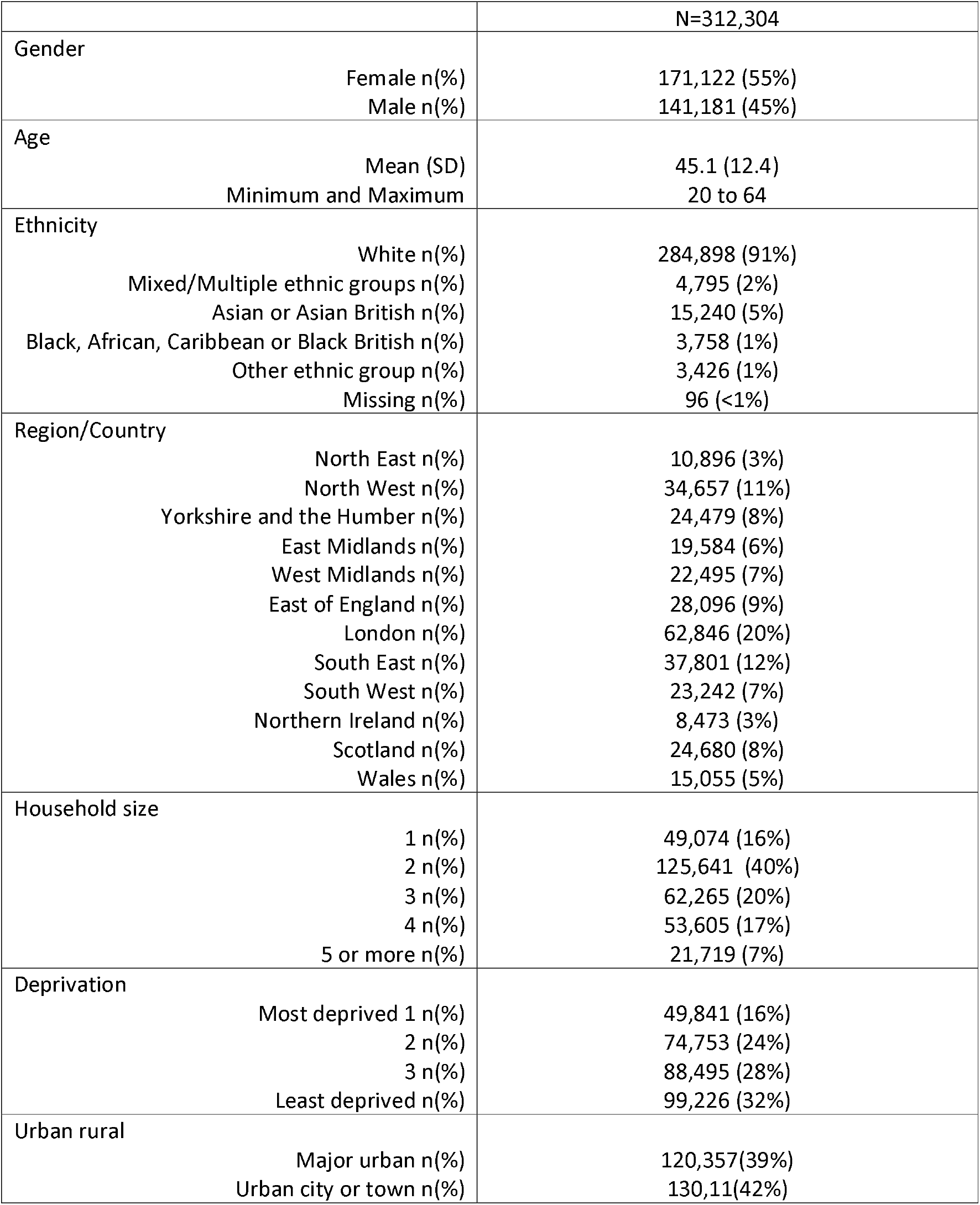
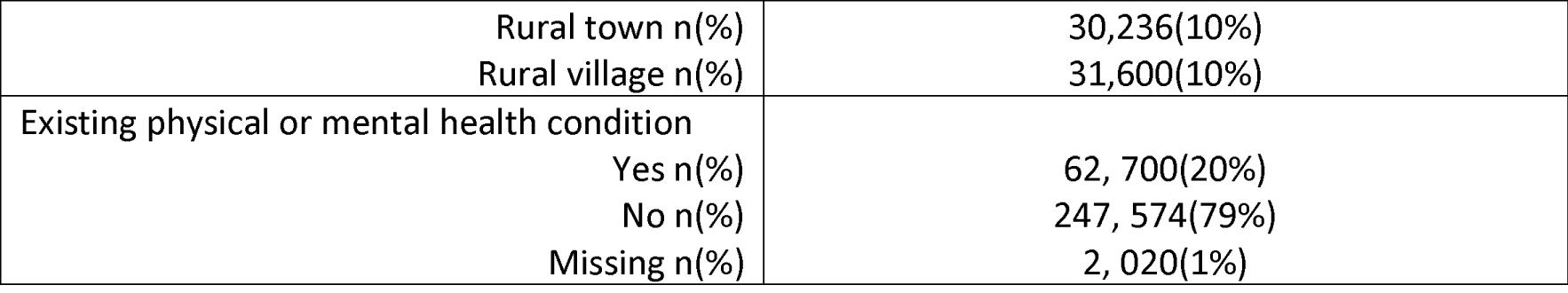
Demographic information of adults aged 20-64

|  |  | N=312,304 |
| --- | --- | --- |
| Gender | Female n(%) | 171,122 (55%) |
|  | Male n(%) | 141,181 (45%) |
| Age | Mean (SD) | 45.1 (12.4) |
|  | Minimum and Maximum | 20 to 64 |
| Ethnicity | White n(%) | 284,898 (91%) |
|  | Mixed/Multiple ethnic groups n(%) | 4,795 (2%) |
|  | Asian or Asian British n(%) | 15,240 (5%) |
|  | Black, African, Caribbean or Black British n(%) | 3,758 (1%) |
|  | Other ethnic group n(%) | 3,426 (1%) |
|  | Missing n(%) | 96 (<1%) |
| Region/Country | North East n(%) | 10,896 (3%) |
|  | North West n(%) | 34,657 (11%) |
|  | Yorkshire and the Humber n(%) | 24,479 (8%) |
|  | East Midlands n(%) | 19,584 (6%) |
|  | West Midlands n(%) | 22,495 (7%) |
|  | East of England n(%) | 28,096 (9%) |
|  | London n(%) | 62,846 (20%) |
|  | South East n(%) | 37,801 (12%) |
|  | South West n(%) | 23,242 (7%) |
|  | Northern Ireland n(%) | 8,473 (3%) |
|  | Scotland n(%) | 24,680 (8%) |
|  | Wales n(%) | 15,055 (5%) |
| Household size | 1 n(%) | 49,074 (16%) |
|  | 2 n(%) | 125,641 (40%) |
|  | 3 n(%) | 62,265 (20%) |
|  | 4 n(%) | 53,605 (17%) |
|  | 5 or more n(%) | 21,719 (7%) |
| Deprivation | Most deprived 1 n(%) | 49,841 (16%) |
|  | 2 n(%) | 74,753 (24%) |
|  | 3 n(%) | 88,495 (28%) |
|  | Least deprived n(%) | 99,226 (32%) |
| Urban rural | Major urban n(%) | 120,357(39%) |
|  | Urban city or town n(%) | 130,11(42%) |

|  |  |
| --- | --- |
| Rural town n(%) | 30,236(10%) |
| Rural village n(%) | 31,600(10%) |
| Existing physical or mental health condition |  |
| Yes n(%) | 62,700(20%) |
| No n(%) | 247,574(79%) |
| Missing n(%) | 2,020(1%) |

Fourdigit Standard Occupational Classification (SOC) 2010 codes were used to derive occupational categorisations. The SOC classification is hierarchical with the first digit indicating major occupational groups, the second, third and fourth digits classifying occupations with increasingly greater detail. Where a participant had a valid two digit SOC recorded but not a four-digit SOC, we utilised their data wherever possible but classified them as missing for other analyses. To allow comparison with other studies we used 13 categories of essential worker defined by Nafilyan et al. (10) and broad categories (for analysis by time period) used by Mutambudzi et al. (8) (See appendix 2). Two additional classifications were one-digit SOC codes and sector (using categories defined by the ONS).

After using available four-digit SOC codes to create the occupational categories, anyone who was recorded as ‘not working’ due to either unemployment, furlough, retirement, childcare, education or other reasons was categorised as ‘not working/student’. Anyone who was working but did not have data on either their SOC code or employment status was recorded as missing. For the cox regressions, work status was time varying. In summary tables and the logistic regression, first available work categorisation (overall or within a time period) was used.

All analyses were conducted in Stata version 17 (25) in the ONS Secure Research Service (SRS).

### Sensitivity analyses

There were several different categorisations of ‘not working’ in the CIS dataset. Data were reanalysed using alternative classifications to see whether these changed conclusions. Due to missing four-digit SOCs for some participants, we ran sensitivity analyses using multiple imputation to impute four-digit SOCs based on two-digit SOCs and demographic information.

We used different definitions of reinfection after the first test in the dataset to see whether these affected results in the analysis by time (using different time gaps and/or numbers of negative tests in between).

We used separate models for North and Midlands vs South of UK. We also used models that included additional variables relating to behaviour outside the workplace and foreign travel.

## Results

By November 2021 there were 312 304 participants of working age in the CIS, of these 25 377 (8%) had at least one infection detected by a PCR test as part of the survey. Table 3 shows demographic information. The group had more females than males (55% vs 45%), and had a mean age of 45 years. Participants contributed information on a total of 3 910 311 visits, with between 1 and 24 visits per person (mean 12.5 visits). A large proportion (91%) classed themselves as part of a White ethnic group and there was some overrepresentation of the least deprived IMD quartile (33% in the 1^st^ quartile compared to 16% in the fourth quartile). 153 302 (49%) were known to be working and had occupational information in the form of a four digit SOC for at least one time point in the survey and 242 904 (78%) had information on industrial sector.

Figure 1 and S1 shows hazard ratios with 95% confidence intervals comparing the hazard of infection for participants in 13 groups of essential worker when compared to non-essential workers. Elevated risks of infection can be seen for social care staff (HR 1.14; 95% CI 1.04 to 1.24), education (HR 1.31; 95% CI 1.23 to 1.39), bus and coach drivers (1.43; 95% CI 1.03 to 1.97) and police and protective services (HR 1.45; 95% CI 1.29 to 1.62) when compared to non-essential workers. It was unclear whether risk of infection was elevated for healthcare support workers 1.13 (95% CI 0.96 to 1.32), food retail and distribution (HR 1.02; 95% CI 0.93 to 1.13), food production (HR 1.04; 95% CI 0.83 to 1.31), taxi and cab drivers and chauffeurs (HR 1.17; 95% CI 0.83 to 1.65), van drivers (HR 1.17; 95% CI 0.92 to 1.23) and other transport workers (HR 1.06; 95% CI 0.92 to 1.23). For healthcare associate professionals (HR 0.96; 95% CI 0.88 to 1.04) there was little evidence of any elevated risk. Healthcare professionals (HR 0.78; 95% CI 0.67 to 0.91) had a small reduction in hazard when compared to non-essential workers during the time period of interest. Adjusting for multiple demographic factors slightly changed the hazards ratios and their confidence intervals.

**Figure 1.**
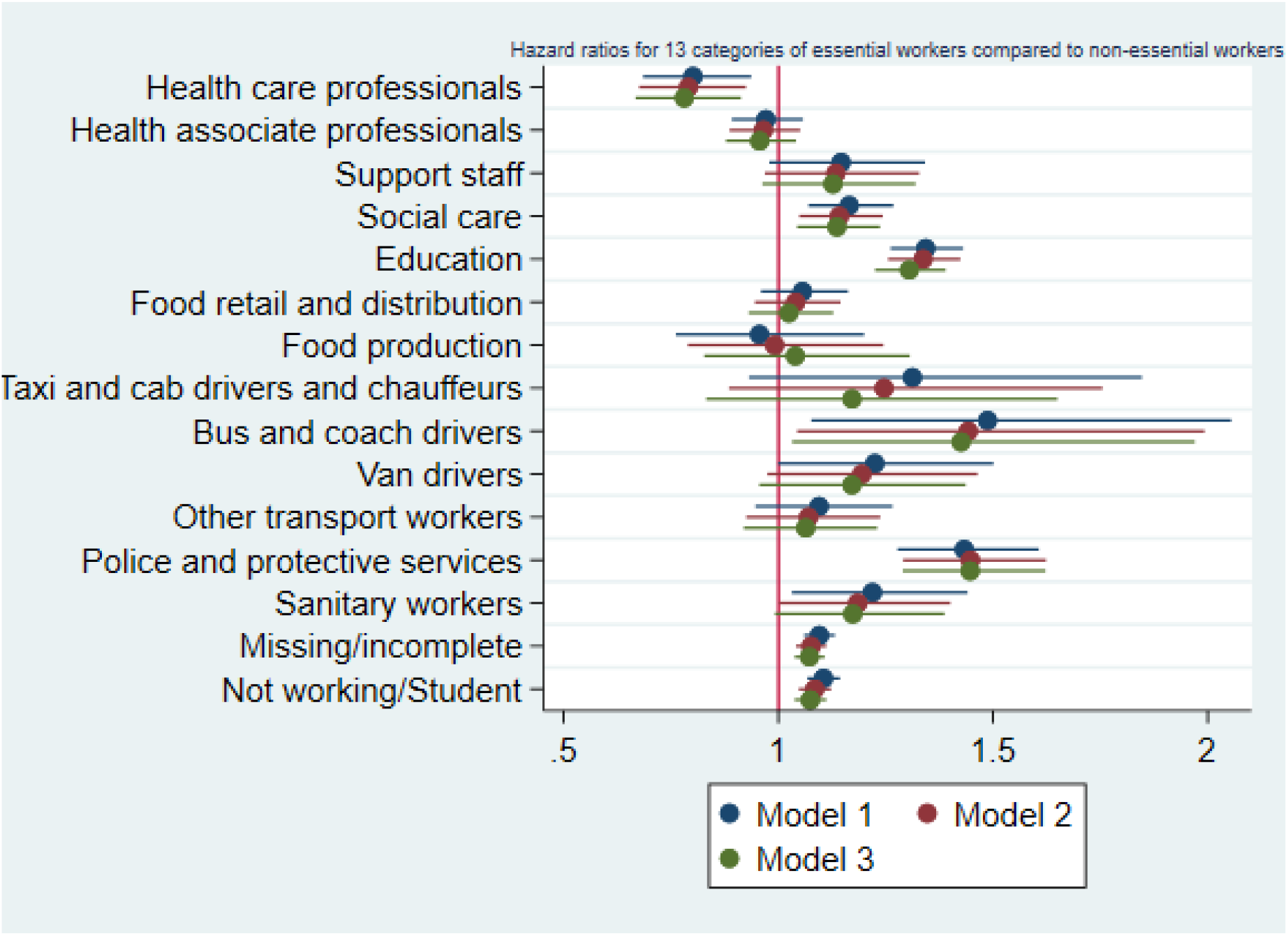
Results of Cox regression for 13 categories of essential worker compared to non-essential workers. Time to first infection adjusted for (1) age and sex (2) age, sex, IMD, ethnic group, region (3) age, sex, IMD, ethnic group, region, household size, rural or urban location, health conditions. Model uses 3772517 observations from 312 304 participants.

When repeating this analysis by sector (S3 and S4), elevated risks on average for the education, social care, food production and transport sectors were observed. The healthcare sector also displayed an increased risk on average. Elevated risks were also observed for retail, hospitality, personal services, financial services, construction, manufacturing and civil service.

Figure 2 shows the interaction between occupation and time for three broad categories of essential workers. There was evidence of an interaction between occupation and time demonstrating that the relative effect of working in an essential occupation on the risk of infection varied over the pandemic. On average healthcare workers had an elevated risk of infection during the earlier periods of the pandemic (April 2020 to September 2020 (T1), October 2020 to February 2021 (T2)), but this diminished by T3 (March 2021 to May 2021, and by T4 (June 2021 to October 2021) was at a level below that of non-essential workers. The combined group of social care and education workers did not have an elevated odds during T1, but did by T2 and this persisted through T3 and T4. For other essential workers on average, elevated risks were seen at T2 and T3, but not at T1 and T4.

**Figure 2:**
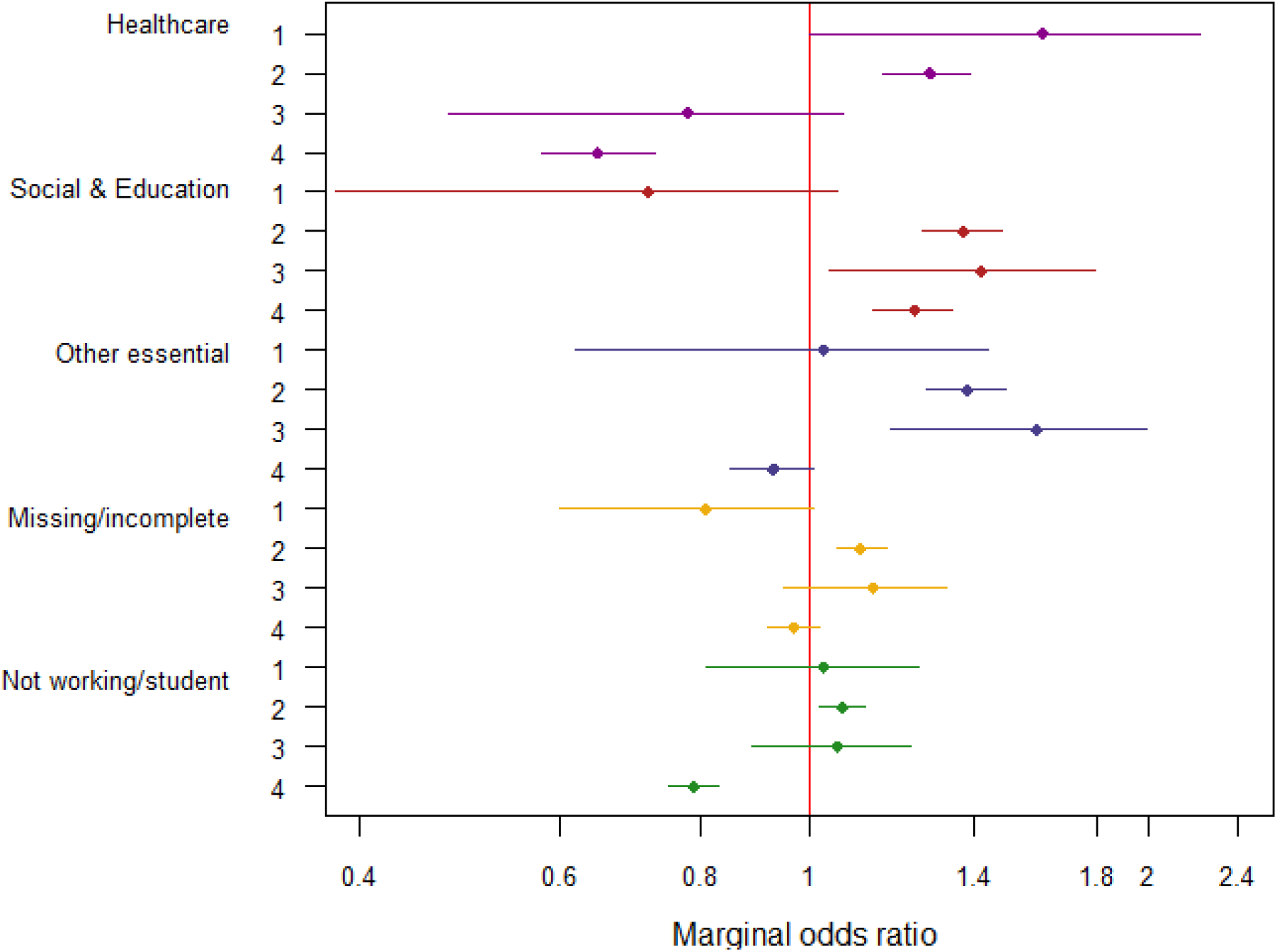
Marginal odds ratios (95% CIs) for odds of new infection for essential worker categories compared to other workers in four time tranches: April 2020 to Sept 2020 (T1), Oct 2020 to Feb 2021 (T2), March 2021 to May 2021 (T3), June 2021 to Oct 2021 (T4). Adjusted for age quintile, sex, ethnicity, IMD, region, household size, urban vs rural area, presence of health conditions. Restricted to working age adults (20-64 years). N= 312 304.

Repeating this analysis by sector for two time periods (S8) shows that for most sectors any elevated odds were most pronounced in the early part of the pandemic, with only education, hospitality and manufacturing having elevated relative odds in both time periods.

The impact of sensitivity analyses on estimates was negligible, and overall conclusions did not change.

## Discussion

There was clear evidence that the relative effect of working in an occupational group varied over the pandemic. During April 2020 to February 2021 when a large number of restrictions were in place, healthcare workers saw an elevated risk compared to non-essential workers. Later, during from March to November 2021 once restrictions were eased, and healthcare workers were offered early vaccines, healthcare did not appear to show an elevated risk compared to other workers. For workers in social care and education, there was little evidence of increased risk in April to September 2020 when schools were mainly closed, but there was a large increase subsequently and this persisted over the time period covered in this analyses. Similar results were observed in another UK cohort, Virus Watch (29) which found a persistently elevated risk for teachers in the third wave of the pandemic compared to other occupations.

We saw elevated risks for most types of workers whose attendance at the workplace was expected during the pandemic. Elevated risks for healthcare workers varied by type and appeared to be only evident early in the pandemic. Note that the infection survey did not start until April 2020 and therefore during the period of study many health care workers may have had immunity from prior infection. In the UK workers in health and social care were offered vaccines earlier than other occupational groups. The fact that healthcare associates and healthcare support staff saw increased risks where healthcare professionals did not is in line with other research (26, 27). Healthcare support workers and healthcare associate professionals would have had similar access to vaccines; but it has been reported that access to PPE for healthcare staff varies by role and work area (28). Some health associate professionals such as dentists and opticians had less face-to-face contact with patients during the first national lockdown and therefore less early exposure.

Food production workers did not see an elevated risk when compared to non-essential workers; seen in both the analysis by sector and by essential worker groups and mirroring analyses of COVID-19 mortality (16). The food production industry has reported a large number of outbreaks with one UK study (18) reporting that this sector had more reported outbreaks per facility than any other sector. The defined group in this study combined outdoor agricultural workers with indoor process operatives; therefore, it may be that the average result disguises heterogeneity of risks within the sector. In addition, migrant and temporary workers, thought to be common in this sector, may be missing from the CIS. Food processing workers may have immunity from early infection and/or high levels of PPE and other mitigation.

We observed increased risks on average for education workers which became evident late in 2020 and persisted over time; this group has been shown to have an above average level of exposure with a high number of close contacts and a high probability of intense space sharing during the working day (29).

Elevated risks were seen for the transport sector overall; small numbers lead to wide confidence intervals when we look at individual groups of transport workers, so the picture as to which groups are most at risk is still unclear.

Several sectors and occupations saw elevated risks in the earlier time periods which diminished later. It is possible that with reduced national restrictions and increased social mixing, that differences in workplace transmission have become less pronounced because the main routes of transmission are now outside the workplace. It is also possible that the differences are to do with increased transmission in the references categories used rather than relative reductions in the categories perceived to be at high risk.

As can be seen in Figure 1 adjustment for other variables made very little difference to our estimates in contrast to analyses of mortality (16). Our results appear robust, regardless of the chosen DAG and adjustment set.

The fact that occupational differences in the rate of COVID-19 exist, even after adjustment for other variables has clear implications for policy and practice. Some sectors and occupations appear to have persistent high risks even after taking into account confounding factors such as age and comorbidity and therefore workplaces and governments need to invest in mitigation measures and further research into how to reduce these risks.

### Strengths

For these analyses, we utilised Sars-Cov-2 infection identified via PCR tests during a longitudinal prevalence survey. These tests would identify both symptomatic and asymptomatic infections. This dataset is ideal for assessing questions about occupation and COVID-19 infection because PCR results via survey visits are likely to be independent of occupation, in contrast to self-reported test results when different occupations would have different access to testing and motivation for testing (30).

We used both occupational groups categorised from 4-digit SOC codes and sector groupings from self-reported sector categories and conclusions remained similar for each. We have used occupational groupings used in previous studies to allow triangulation of results.

We looked at how the relative risks between occupations changed over time; these analyses both allowed us to see how relative effects changed according to the set of restrictions and mitigation strategies in place at the time. They also allowed us to take into account that for some workers in the UK either their occupation or working status changed over time, as well as being able to include reinfections for the same individual.

### Limitations

The ONS infection survey was a prevalence study which in the most part conducted monthly tests for participants; it is likely that positive results were missed in between visits. While this is relevant to prevalence estimates, it is less likely to affect relative effects. The CIS started in April 2020, several months into the pandemic, so we are likely to have missed a period where occupational risks would have been most evident for some occupational groups.

It is possible that certain occupations will be underrepresented in the survey due to their availability for the study visits because of long working hours or shift work. There is risk of selection bias; e.g. healthcare workers who were front-line may have been less likely to be recruited or less likely to provide data than those who were non-frontline due to shift work or lack of time.

Occupational information, particularly 4-digit SOC, was missing for a proportion of participants. We used sensitivity analyses and also accompanied analyses by occupation with analysis by sector (where information was more complete) in an attempt to check that our findings are robust.

There is likely to be variation within occupational groups and sectors that will be masked when assessing group averages. The sample is not large enough to make meaningful analysis of more granular groupings, particularly when assessing separate time periods.

## Conclusions

Some occupational groups see elevated risks of Covid-19 infection when compared to others, and the relative effects varied at different time-points during the pandemic. Increased risks for healthcare workers appear to be most pronounced during the early part of the pandemic, but varied according to the type of healthcare worker. Increased risks were seen for workers in the education and social care sectors once the initial lockdown of the first wave was over, and this has persisted into the third wave suggesting that increased mitigation is required in these sectors.

## Supporting information

Strobe checklist

## Data Availability

This data can be accessed only by researchers who are Office of National Statistics (ONS) accredited researchers. 
Researchers can apply for accreditation through the Research Accreditation Service. Access is through the Secure Research Service (SRS) and approved on a project basis. For further details see
https://www.ons.gov.uk/aboutus/whatwedo/statistics/requestingstatistics/approvedresearcherscheme

## Funding

This work was supported by funding through the National Core Study “PROTECT” programme, managed by the Health and Safety Executive on behalf of HM Government. SVK acknowledges funding from a NRS Senior Clinical Fellowship (SCAF/15/02), the Medical Research Council (MC_UU_00022/2) and the Scottish Government Chief Scientist Office (SPHSU17).

## Declaration

This work was produced using statistical data from ONS. The use of the ONS statistical data in this work does not imply the endorsement of the ONS in relation to the interpretation or analysis of the statistical data. This work uses research datasets which may not exactly reproduce National Statistics aggregates

## Research Ethics Approval

UK Statistics Authority self assessment classified study as low risk. This assessment was verified by the Office for National Statistics (ONS) Research Accreditation Panel.

## Notes

### Competing Interest Statement

All authors have completed the ICMJE uniform disclosure form at www.icmje.org/coi_disclosure.pdf and declare: SR, JW, NP, WM, MC, KS, MG, MVT had funding for this work through the National Core Study PROTECT programme, managed by the Health and Safety Executive on behalf of HM Government. SVK acknowledges funding from a NRS Senior Clinical Fellowship (SCAF/15/02), the Medical Research Council (MC_UU_00022/2) and the Scottish Government Chief Scientist Office (SPHSU17). KS reports funding from the NIHR via a Doctoral Research Fellowship.

### Author Declarations

The COVID-19 Infection Survey (CIS) has been given approval by South Central Berkshire B Research Ethics Committee (20/SC/0195). Office for National Statistics (ONS) Research Accreditation Panel verified UK Statistics Authority self assessment for secondary data analysis (study classified as low risk).

### Summary of Updates

Corrections to the links to tables and figures

