## Supplementary materials for "Occupational differences in SARS-CoV-2 infection: Analysis of the UK ONS Coronavirus (COVID-19) Infection Survey"

**Appendix 1. Supplementary materials.**

This work was produced using statistical data from ONS. The use of the ONS statistical data in this work does not imply the endorsement of the ONS in relation to the interpretation or analysis of the statistical data. This work uses research datasets which may not exactly reproduce National Statistics aggregates

S1 Hazard ratios and 95% CI from time-varying Cox regression with 13 categories of essential worker compared to non-essential workers. Time to first infection adjusted for (1)age and sex (2)age, sex, IMD, ethnic group, region (3) age, sex, IMD, ethnic group, region, household size, rural or urban location, health conditions. Model uses 3772517 observations from 312 304 participants.

| Model | (1) | (2) | (3) |
| --- | --- | --- | --- |
| Health care professionals | 0.801 [0.685,0.937] | 0.790 [0.675,0.924] | 0.781 [0.667,0.913] |
| Health associate professionals | 0.971 [0.892,1.057] | 0.965 [0.886,1.051] | 0.956 [0.878,1.041] |
| Support staff | 1.146 [0.979,1.342] | 1.134 [0.968,1.328] | 1.127 [0.962,1.320] |
| Social care | 1.165 [1.071,1.268] | 1.142 [1.049,1.243] | 1.136 [1.044,1.237] |
| Education | 1.343 [1.261,1.430] | 1.337 [1.256,1.424] | 1.305 [1.225,1.390] |
| Food retail and distribution | 1.056 [0.960,1.162] | 1.040 [0.945,1.145] | 1.024 [0.931,1.128] |
| Food production | 0.956 [0.761,1.200] | 0.991 [0.789,1.245] | 1.039 [0.827,1.306] |
| Taxi and cab drivers and chauffeurs | 1.312 [0.932,1.847] | 1.246 [0.885,1.755] | 1.171 [0.832,1.650] |
| Bus and coach drivers | 1.487 [1.077,2.055] | 1.442 [1.044,1.993] | 1.426 [1.032,1.970] |
| Van drivers | 1.225 [0.999,1.501] | 1.194 [0.974,1.464] | 1.171 [0.955,1.436] |
| Other transport workers | 1.095 [0.947,1.266] | 1.070 [0.925,1.237] | 1.064 [0.920,1.230] |
| Police and protective services | 1.433 [1.277,1.607] | 1.447 [1.290,1.623] | 1.447 [1.290,1.622] |
| Sanitary workers | 1.219 [1.031,1.441] | 1.185 [1.002,1.401] | 1.173 [0.992,1.387] |
| Non-essential workers | 1 [1,1] | 1 [1,1] | 1 [1,1] |
| Missing/incomplete | 1.095 [1.060,1.132] | 1.076 [1.041,1.112] | 1.072 [1.038,1.108] |
| Not working/Student | 1.105 [1.068,1.144] | 1.085 [1.048,1.123] | 1.074 [1.038,1.112] |

S2 N(%) or participants with at least one positive PCR in the CIS by occupational group

| Occupation | Positive PCR in CIs | N |
| --- | --- | --- |
|  | n(%) |  |
| Health care professionals | 179(6.3) | 2864 |
| Health associate professionals | 685(7.7) | 8874 |
| Support staff | 194(8.7) | 2217 |
| Social care | 690(8.6) | 8005 |
| Education | 1285(10.3) | 12492 |
| Food retail and distribution | 571(8.2) | 6953 |
| Food production | 83(7.4) | 1128 |
| Taxi and cab drivers and chauffeurs | 43(9.3) | 465 |
| Bus and coach drivers | 40(9.9) | 404 |
| Van drivers | 113(9.3) | 1211 |
| Other transport workers | 215(8.6) | 2488 |
| Police and protective services | 322(11.3) | 2852 |
| Sanitary workers | 183(9.4) | 1954 |
| Non-essential workers | 9033(7.8) | 115385 |
| Missing/incomplete | 7001(9.3) | 75706 |
| Not working/Student | 4740(6.8) | 69333 |

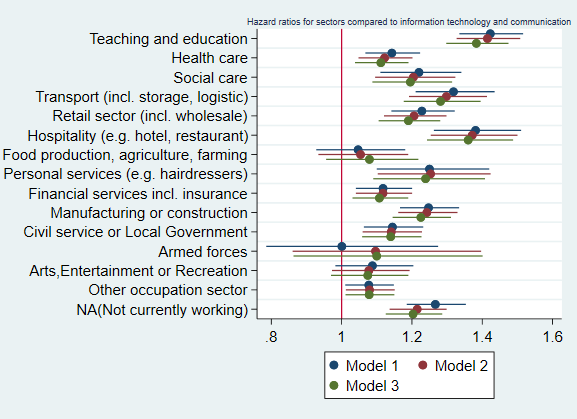

S3. Results of Cox regression by sector, with each sector compared to ‘information technology and communication’ using Hazard ratios. Time to first infection adjusted for (1)age and sex (2)age, sex, IMD, ethnic group, region (3) age, sex, IMD, ethnic group, region, household size, rural or urban location, health conditions. Model uses 3772517 observations from 312 304 participants.

S4. Hazard ratios and 95% CI for time varying Cox regression by sector, with each sector compared to ‘information technology and communication’ using Hazard ratios. Time to first infection adjusted for (1)age and sex (2)age, sex, IMD, ethnic group, region (3) age, sex, IMD, ethnic group, region, household size, rural or urban location, health conditions. Model uses 3772517 observations from 312 304 participants.

|  | (1) | (2) | (3) |
| --- | --- | --- | --- |
| Teaching and education | 1.423 [1.335,1.516] | 1.415 [1.327,1.508] | 1.383 [1.298,1.475] |
| Health care | 1.143 [1.068,1.223] | 1.122 [1.048,1.202] | 1.112 [1.038,1.190] |
| Social care | 1.220 [1.110,1.340] | 1.204 [1.095,1.324] | 1.195 [1.087,1.314] |
| Transport (incl. storage, logistic) | 1.318 [1.211,1.435] | 1.298 [1.192,1.413] | 1.281 [1.177,1.395] |
| Retail sector (incl. wholesale) | 1.228 [1.141,1.322] | 1.206 [1.120,1.298] | 1.190 [1.105,1.281] |
| Hospitality (e.g. hotel, restaurant) | 1.381 [1.262,1.510] | 1.371 [1.253,1.500] | 1.360 [1.243,1.488] |
| Food production, agriculture, farming | 1.047 [0.928,1.181] | 1.054 [0.934,1.189] | 1.079 [0.956,1.218] |
| Personal services (e.g. hairdressers) | 1.250 [1.100,1.420] | 1.253 [1.103,1.424] | 1.239 [1.090,1.408] |
| Information technology and communication | 1 [1,1] | 1 [1,1] | 1 [1,1] |
| Financial services incl. insurance | 1.118 [1.041,1.200] | 1.118 [1.041,1.200] | 1.107 [1.031,1.189] |
| Manufacturing or construction | 1.247 [1.166,1.334] | 1.242 [1.161,1.329] | 1.225 [1.145,1.311] |
| Civil service or Local Government | 1.145 [1.063,1.232] | 1.141 [1.060,1.229] | 1.140 [1.058,1.227] |
| Armed forces | 1.001 [0.786,1.274] | 1.096 [0.861,1.397] | 1.100 [0.863,1.401] |
| Arts,Entertainment or Recreation | 1.088 [0.982,1.204] | 1.077 [0.973,1.193] | 1.074 [0.970,1.190] |
| Other occupation sector | 1.077 [1.010,1.149] | 1.079 [1.012,1.151] | 1.078 [1.011,1.150] |
| NA(Not currently working) | 1.267 [1.185,1.353] | 1.215 [1.137,1.298] | 1.203 [1.125,1.286] |

S5 Hazard ratios and 95% CI for Cox regression with categories by one-digit SOC compared to Managers, directors and senior officials. Time to first infection adjusted for (1)age and sex (2)age, sex, IMD, ethnic group, region (3) age, sex, IMD, ethnic group, region, household size, rural or urban location, health conditions. Model uses 3772517 observations from 312 304 participants.

| Model | (1) | (2) | (3) |
| --- | --- | --- | --- |
| Managers, directors and senior officials | 1 [1,1] | 1 [1,1] | 1 [1,1] |
| Professional occupations | 0.987 [0.936,1.042] | 0.984 [0.932,1.038] | 0.988 [0.936,1.043] |
| Associate professional and technical | 1.004 [0.947,1.065] | 0.998 [0.942,1.058] | 1.010 [0.953,1.071] |
| Admin and secretarial | 1.015 [0.952,1.082] | 1.006 [0.943,1.072] | 1.010 [0.948,1.077] |
| Skilled trades | 1.030 [0.955,1.110] | 1.032 [0.958,1.113] | 1.025 [0.951,1.105] |
| Caring, leisure and other service | 1.330 [1.240,1.427] | 1.311 [1.222,1.407] | 1.290 [1.201,1.384] |
| Sales and customer service | 1.056 [0.969,1.151] | 1.031 [0.946,1.124] | 1.026 [0.941,1.119] |
| Process plant and machine operatives | 1.221 [1.116,1.335] | 1.191 [1.089,1.303] | 1.180 [1.079,1.292] |
| Elementary | 1.204 [1.107,1.309] | 1.175 [1.081,1.279] | 1.163 [1.070,1.265] |
| Not working/Student | 1.039 [0.985,1.095] | 1.022 [0.969,1.077] | 1.029 [0.975,1.085] |
| Missing/incomplete | 0.796 [0.748,0.846] | 0.787 [0.740,0.836] | 0.795 [0.748,0.846] |

S6: New positive PCR tests for essential worker categories compared four time periods: April 2020 to Sept 2020 (T1), Oct 2020 to Feb 2021 (T2), March 2021 to May 2021 (T3), June 2021 to Oct 2021 (T4).

|  | T1 | | T2 | | T3 | | T4 | |
| --- | --- | --- | --- | --- | --- | --- | --- | --- |
|  | Infection | Total | Infection | Total | Infection | Total | Infection | Total |
| Health n | 32 | 4,170 | 577 | 9,698 | 29 | 9,728 | 337 | 11,854 |
| % | 0.77 | 100 | 5.95 | 100 | 0.3 | 100 | 2.84 | 100 |
| Social and Education n | 19 | 5,518 | 795 | 12,501 | 68 | 12,498 | 806 | 15,370 |
| % | 0.34 | 100 | 6.36 | 100 | 0.54 | 100 | 5.24 | 100 |
| Other Essential n | 29 | 5,900 | 828 | 12,945 | 74 | 12,156 | 621 | 15,580 |
| % | 0.49 | 100 | 6.4 | 100 | 0.61 | 100 | 3.99 | 100 |
| Other workers n | 171 | 35,812 | 3,834 | 81,166 | 306 | 79,634 | 4,208 | 98,425 |
| % | 0.48 | 100 | 4.72 | 100 | 0.38 | 100 | 4.28 | 100 |
| Missing/incomplete n | 93 | 24,150 | 3,104 | 59,479 | 265 | 60,755 | 2,544 | 61,191 |
| % | 0.39 | 100 | 5.22 | 100 | 0.44 | 100 | 4.16 | 100 |
| Not working/Student n | 166 | 33,870 | 4,004 | 79,535 | 300 | 73,359 | 2,523 | 74,328 |
| % | 0.49 | 100 | 5.03 | 100 | 0.41 | 100 | 3.39 | 100 |
| Total n | 510 | 109,420 | 13,142 | 255,324 | 1,042 | 248,130 | 11,039 | 276,748 |
| % | 0.47 | 100 | 5.15 | 100 | 0.42 | 100 | 3.99 | 100 |

S7: Marginal odds ratios (95% CIs) for odds of new infection for essential worker categories compared to other workers in four time periods: April 2020 to Sept 2020 (T1), Oct 2020 to Feb 2021 (T2), March 2021 to May 2021 (T3), June 2021 to Oct 2021 (T4). Adjusted for age quintile, sex, ethnicity, IMD, region, household size, urban vs rural area. Restricted to working age adults (20-64 years). N= 312304 participants.

| Work category | Time Tranche | MOR | 95% CI | |
| --- | --- | --- | --- | --- |
| healthcare | 1 | 1.58 | 0.96 | 2.19 |
| healthcare | 2 | 1.29 | 1.18 | 1.41 |
| healthcare | 3 | 0.79 | 0.48 | 1.1 |
| healthcare | 4 | 0.66 | 0.58 | 0.73 |
| social essential | 1 | 0.85 | 0.47 | 1.24 |
| social essential | 2 | 1.38 | 1.27 | 1.48 |
| social essential | 3 | 1.53 | 1.14 | 1.93 |
| social essential | 4 | 1.21 | 1.11 | 1.3 |
| Other essential | 1 | 1.08 | 0.66 | 1.49 |
| Other essential | 2 | 1.32 | 1.22 | 1.43 |
| Other essential | 3 | 1.56 | 1.16 | 1.96 |
| Other essential | 4 | 0.95 | 0.86 | 1.03 |
| missing/incomplete | 1 | 1.51 | 1.15 | 1.87 |
| missing/incomplete | 2 | 1.04 | 0.997 | 1.09 |
| missing/incomplete | 3 | 1.16 | 0.97 | 1.35 |
| missing/incomplete | 4 | 1.08 | 1.03 | 1.14 |
| not working/student | 1 | 1.06 | 0.83 | 1.29 |
| not working/student | 2 | 1.06 | 1.01 | 1.11 |
| not working/student | 3 | 1.09 | 0.92 | 1.27 |
| not working/student | 4 | 0.81 | 0.77 | 0.85 |

S8 Marginal odds ratios from multilevel logistic regression based on first available sector group, and any new infection within the time tranche. Reference category non-essential workers. Two time tranches: T1 1st April 2020 – 28th Feb 2021 and T2 1st March 2021 – 31st Oct 2021. Models based on 312 304 participants. Analysis adjusted for sex, age, region, ethnic group, IMD, rural or urban location, household size, health. N= 312 304 participants.

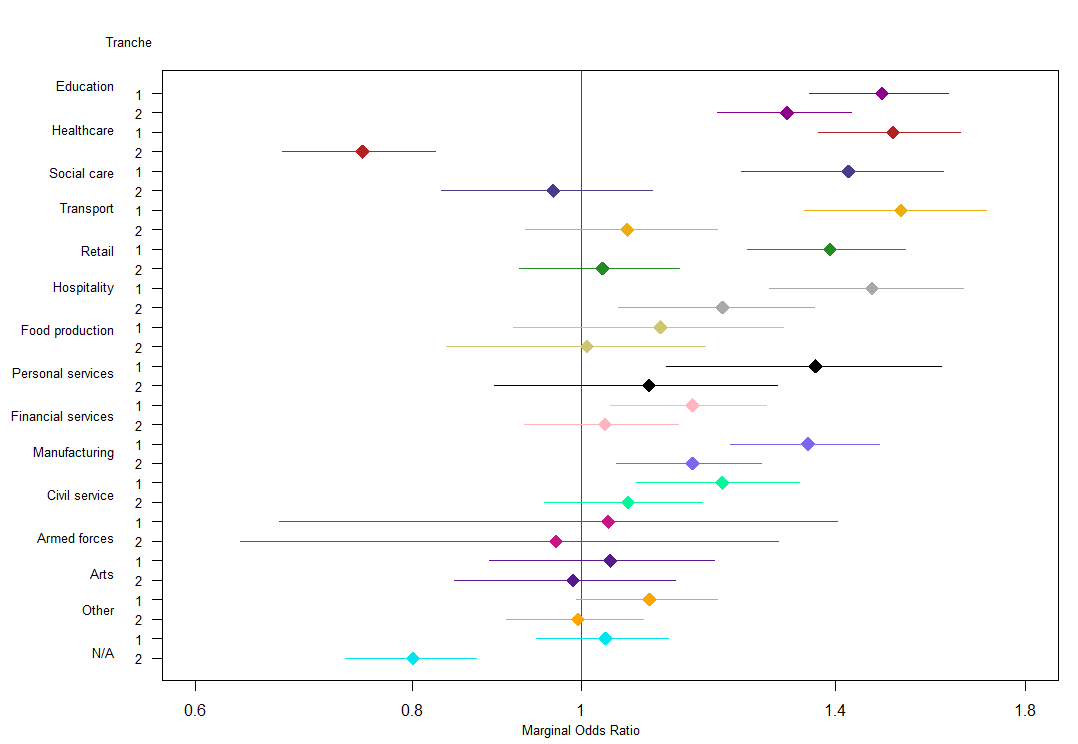

S9: Directed acyclic graph for the relationship between occupation and COVID-19.

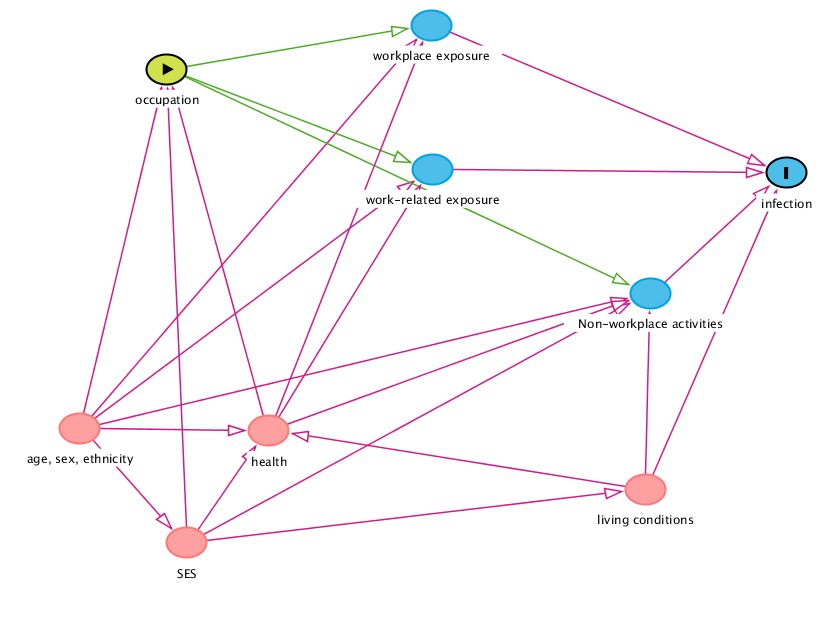
